# Conditions of Confinement in U.S. Carceral Facilities during COVID-19: Individuals Speak: Incarcerated during the COVID-19 Epidemic (INSIDE)

**DOI:** 10.1101/2022.03.15.22271255

**Authors:** Nicole Cassarino, Harika Dabbara, Carla B. Monteiro, Arthur Bembury, Leslie Credle, Uma Grandhi, Samantha White, Monik C. Jiménez

**Author notes:** Both authors contributed equally to the preparation of the manuscript and are listed in alphabetical order (Nicole Cassarino, Harika Dabbara). **Corresponding Author:** Monik C. Jiménez, ScD, SM, FAHA, Division of Women’s Health, Brigham and Women’s Hospital, Harvard Medical School, 1620 Tremont St., 3-034, Boston, MA 02120. **Author contributions:** MCJ initiated the study and led framing of the study, contributed to the analysis and drafted the manuscript; AB, CM, HD, LC and NC conceptualized the study design and contributed to drafting the manuscript; NC led and conducted the data analysis; HD led manuscript preparation; HD and NC jointly led data interpretation. AB, CM, HD, LC, MJ, NC, SW and UG contributed to interpreting the results and all approved the final version of the submitted manuscript.

## Abstract

**Objectives:** We aimed to describe conditions of confinement among people incarcerated in the United States during the coronavirus disease 2019 (COVID-19) pandemic and assess the feasibility of a community-science data collection approach.

**Methods:** We developed a web-based survey with community partners to collect information on confinement conditions (COVID-19 safety, basic needs, support). Formerly incarcerated adults released after March 1, 2020, or non-incarcerated adults in communication with an incarcerated person (proxy) were recruited through social media from July 25, 2020, through March 27, 2021. Descriptive statistics were estimated in aggregate and separately by proxy or formerly incarcerated status. Additionally, we compared responses between proxy and formerly incarcerated respondents using chi-square or Fisher’s exact tests as appropriate based on alpha=0.05.

**Results:** Of 378 responses, 94% were by proxy, and 76% reflected state prison conditions. Participants reported inability to physically distance (≥6ft at all times) (92%), inadequate access to soap (89%), water (46%), toilet paper (49%) and showers (68%). Among people who received mental healthcare before the pandemic, 75% reported reduced care. We found that responses were consistent between formerly incarcerated people and proxy-respondents.

**Conclusions:** Our findings suggest that a community-science approach to data collection is feasible. Based on these findings, COVID-19 safety and basic needs were not sufficiently addressed within some carceral settings. Thus, we recommend the lived experiences of incarcerated individuals should be included to make informed and equitable policy decisions.

## INTRODUCTION

Over a year after the initial global response to coronavirus disease 2019 (COVID-19), the United States (U.S.) sits at the top of two lists: the highest rate of incarceration, and the highest rate of COVID-19 deaths.^1,2^ In 2020, COVID-19 case rates among individuals in U.S. federal and state departments of corrections (DOC) were 4.8-5.5 times higher than the general population.^3-5^ Previous outbreaks of influenza,^6-8^ Legionnaires’ disease,^9^ H1N1,^10,11^ and tuberculosis^12^ were harbingers of carceral facilities’ vulnerability to SARS-CoV-2. Conditions such as overcrowding, inadequate ventilation, poor sanitation, and difficulty accessing personal protective equipment (PPE) have created environments primed for outbreaks. Additionally, highly prevalent conditions like diabetes, hypertension, and asthma place incarcerated populations at greater risk of COVID-19-related morbidity and mortality.^13^

Physical distancing, mask wearing, handwashing, and testing have been key public health COVID-19 mitigation strategies.^14^ However, anecdotal reports indicate that mitigation strategies reported by state-level DOCs are irregularly implemented and highly punitive. Incarcerated individuals report inadequate access to soap, water, and disinfectant supplies,^15^ and extensive periods of lockdowns (suspension of activities and confinement restricted to housing areas) or solitary confinement (housing with minimal to rare contact with others) to achieve physical distancing.^15-17^ The punitive nature of these measures may have unknown consequences for physical and mental health.^18^

While anecdotal evidence is compelling, robust data on the lived experience of incarcerated people during the pandemic is limited with only one quantitative peer reviewed study to date.^19^ Moreover, participants were only included from prisons in three Midwestern or Southeastern states. Due to the decentralized response to COVID-19 across state carceral systems, data from more states and facility types are needed to understand the lived experience of incarceration during the COVID-19 pandemic.

Therefore, we partnered with community advocates to design and disseminate a web-based survey to collect detailed data on the conditions of confinement and determine the feasibility of collecting data through outreach to community members such as loved ones, formerly incarcerated individuals (FII), and professional partners.

## METHODS

### Instrument Development

In May 2020, a Community Advisory Board (CAB) was convened to design a survey to examine condition experienced by individuals incarcerated during the pandemic. The CAB included FII advocates from local and national organizations, Families for Justice as Healing, Justice 4 Housing, the National Council of Incarcerated and Formerly Incarcerated Women and Girls, and the Partakers Organization. Each member described their commitment to their communities below.

*“My work and experience with the criminal justice system has been the impetus that motivates me to seek solutions that can help reduce recidivism by providing essential resources to formerly incarcerated individuals* … *I believe in working with the gatekeepers to ensure that funding is directed to critical areas that are needed most, and will have a significant impact on the population and the communities we work with.”* —Arthur Bembury

*“Because of the things I saw during my 19 years of incarceration I have become an advocate for the women I had to leave behind and the women that will sleep in a prison bed tomorrow.”* —CAB Member

*“Surviving the Federal carceral punishment system is the motivation that drives the work I do today. At Justice 4 Housing we advocate for the abolishment of discriminatory housing policies and ending incarceration of women.”* —Leslie Credle

At study onset, there were no validated instruments to examine confinement conditions among incarcerated individuals during the pandemic; therefore, we developed a novel survey across four domains: COVID-19 safety, basic needs, support, and demographics factors. Each domain was developed based on carceral system mitigation strategies, Center for Disease Control and Prevention (CDC) guidelines,^14^ community-informed concerns of CAB members,^20^ publicly available anecdotal evidence, and surveys targeting the general population.

The instrument was piloted among twelve individuals who had been incarcerated and released after March 1, 2020, or were in communication with an incarcerated individual. The pilot survey included questions regarding estimated time of completion, accessibility, necessity of questions asked, and inclusion of additional questions to better capture lived experiences. In partnership we reviewed the pilot findings, implemented changes, and planned survey dissemination.

The final instrument consisted of 42 questions, (37 multiple choice and 5 open response) (Online Supplement), of which three were administered only to proxies and seven only to FII. Data were collected anonymously using REDCap and the survey was distributed using a snowball sampling method through Twitter and Facebook and by email to community-partner organizations, medical and legal professionals, and local politicians. After deployment, proxy respondents were allowed to indicate their medical or legal professional status.

### Study Sample

Participants were required to be aged ≥18 years and either (1) a formerly incarcerated individual (FII), released after March 1, 2020 (beginning of the U.S. response to the pandemic), or (2) in contact with someone currently incarcerated (proxy). Currently incarcerated individuals were not recruited. To maximize reported data questions could be skipped. Proxies in contact with multiple incarcerated individuals were asked to submit a separate survey for each incarcerated person, unless proxies were legal or medical professionals, who were prompted to report the number of people for whom the information was applicable.

## STATISTICAL ANALYSIS

All primary analyses were descriptive and thus did not include statistical hypothesis testing. Frequencies and relative frequencies were calculated for each question based on available data for all respondents in aggregate, with a missing indicator for skipped questions. Geographic regions were defined according to the U.S. Census Bureau classifications.^21^

We assessed internal validity by comparing the distribution of responses (excluding missing values) by proxies to FII respondents for questions regarding COVID-19 safety, basic needs, and support. Comparisons were conducted using two-sided Fisher’s Exact Tests based on an α=0.05. In addition, we conducted sensitivity analyses to examine differences over time and state of incarceration for any variables with inconsistencies between proxy- and FII respondents. All analyses were conducted using Stata/IC (version 16.1, College Station, TX).

## STATEMENT OF ETHICS

This study was approved by the institutional review board of Mass General Brigham and deemed exempt because no personal identifiers were collected, and the research posed minimal risk. All procedures were in accordance with institutional guidelines. However, we acknowledge that these data represent the lived experiences of individuals who have been incarcerated and those who know them personally.

## RESULTS

Of 378 responses collected between July 28, 2020 and March 27, 2021, 94.4% (n=357) were proxies who completed the survey based on experiences shared by someone currently incarcerated (Table 1). The remaining 5.6% (n=21) were FII who were released after March 1, 2020 (Table 1). Most respondents provided data on facilities in the Northeast (37.0%), followed by the West (28.7%), South (26.1%), and Midwest (8.2%). In addition, 75.7% of respondents reflected conditions in state prisons, followed by jails (13.5%), federal prisons (5.8%), and other facilities including Immigration and Customs Enforcement (ICE) and juvenile detention centers (5.0%) (Table 1). Among respondents who reported gender of the impacted person, 39.6% identified as female, 60.1% were White, followed by Black (21.5%), and Hispanic/Latinx (12.9%).

**Table 1.**
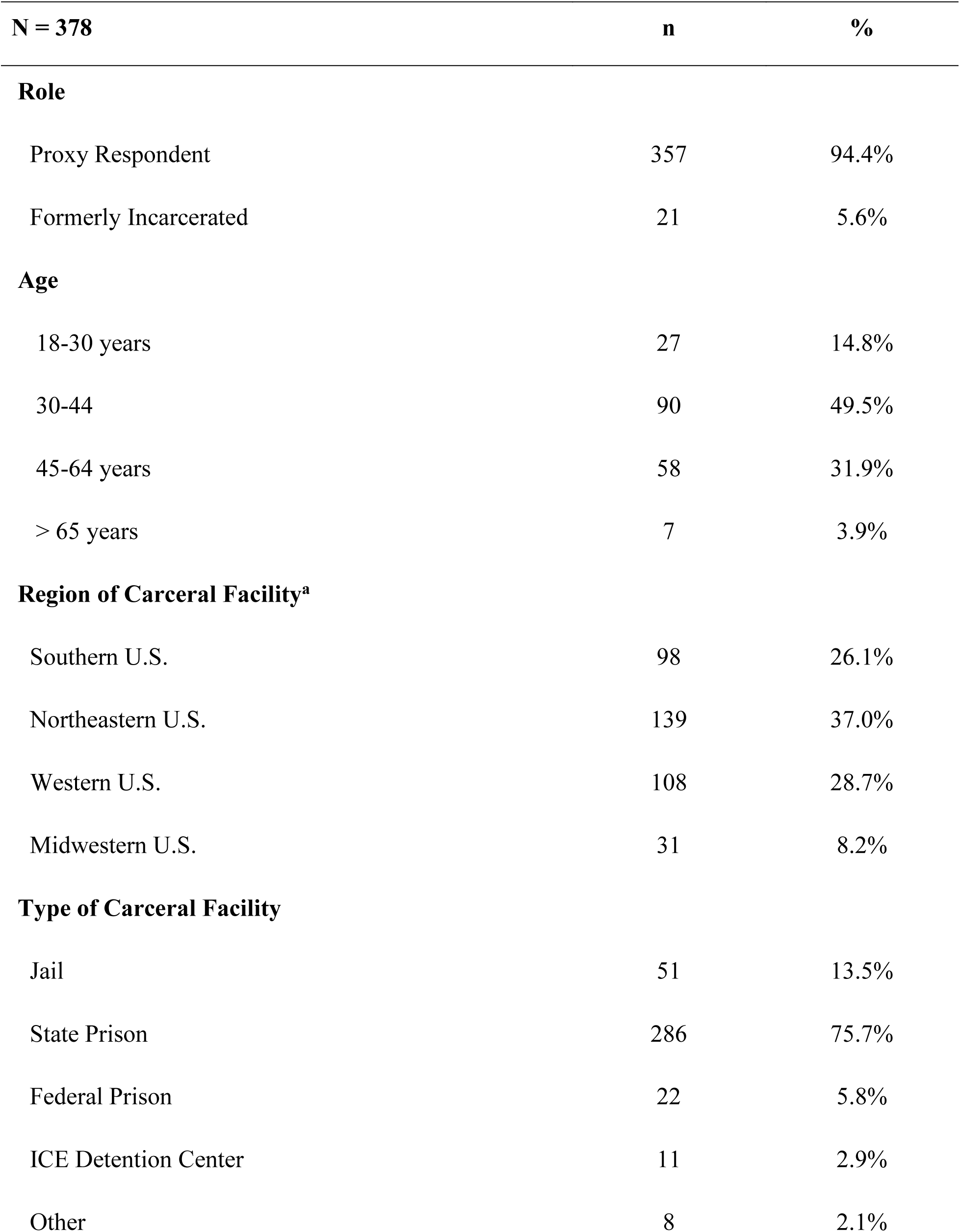
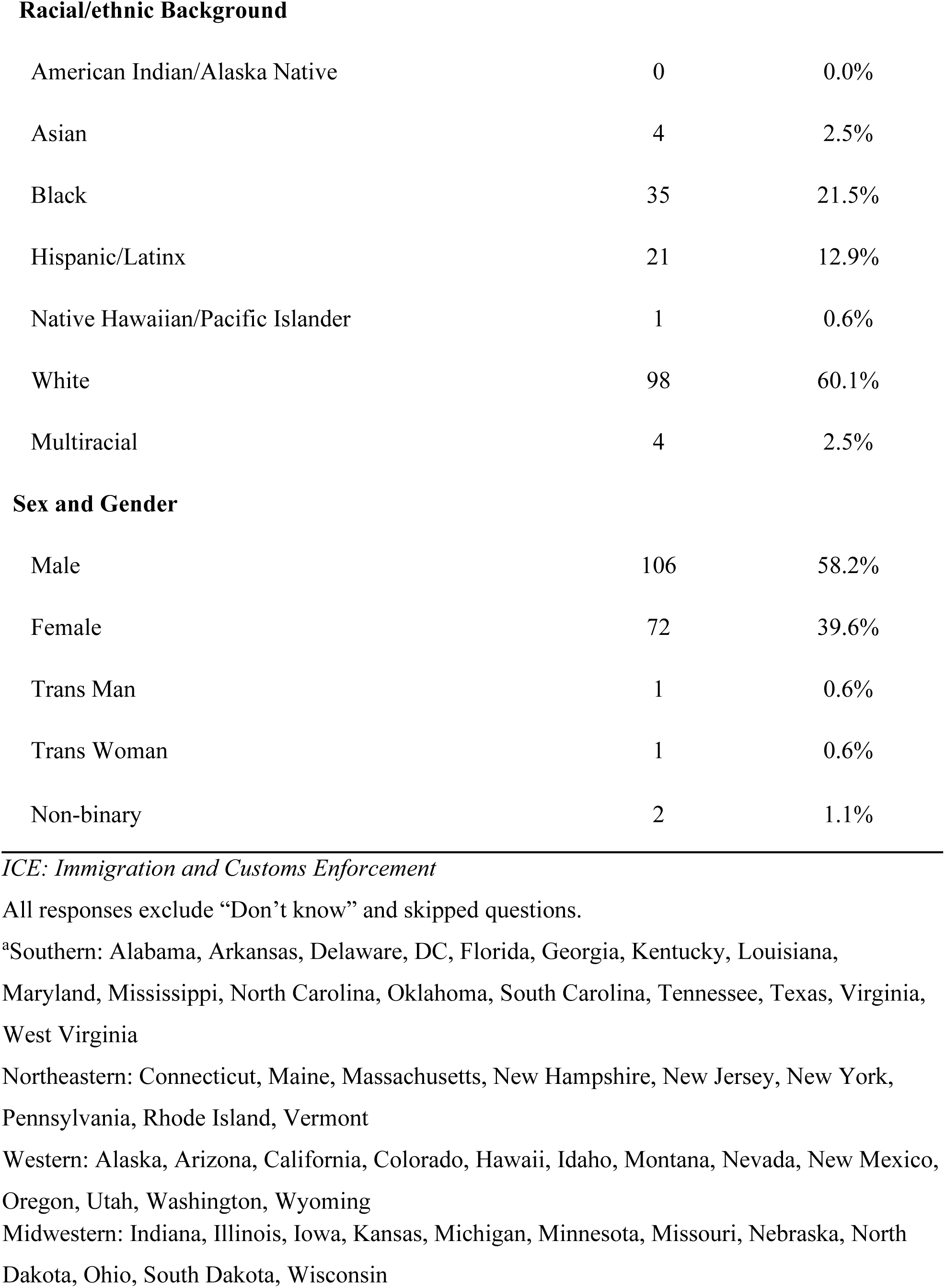
Facility and Demographic Information of Incarcerated People Represented.

| **N = 378** | **n** | **%** |
| --- | --- | --- |
| **Role** |  |  |
| Proxy Respondent | 357 | 94.4% |
| Formerly Incarcerated | 21 | 5.6% |
| **Age** |  |  |
| 18-30 years | 27 | 14.8% |
| 30-44 | 90 | 49.5% |
| 45-64 years | 58 | 31.9% |
| > 65 years | 7 | 3.9% |
| **Region of Carceral Facility<sup>a</sup>** |  |  |
| Southern U.S. | 98 | 26.1% |
| Northeastern U.S. | 139 | 37.0% |
| Western U.S. | 108 | 28.7% |
| Midwestern U.S. | 31 | 8.2% |
| **Type of Carceral Facility** |  |  |
| Jail | 51 | 13.5% |
| State Prison | 286 | 75.7% |
| Federal Prison | 22 | 5.8% |
| ICE Detention Center | 11 | 2.9% |
| Other | 8 | 2.1% |
| **Racial/ethnic Background** |  |  |
| American Indian/Alaska Native | 0 | 0.0% |
| Asian | 4 | 2.5% |
| Black | 35 | 21.5% |
| Hispanic/Latinx | 21 | 12.9% |
| Native Hawaiian/Pacific Islander | 1 | 0.6% |
| White | 98 | 60.1% |
| Multiracial | 4 | 2.5% |
| **Sex and Gender** |  |  |
| Male | 106 | 58.2% |
| Female | 72 | 39.6% |
| Trans Man | 1 | 0.6% |
| Trans Woman | 1 | 0.6% |
| Non-binary | 2 | 1.1% |

*ICE: Immigration and Customs Enforcement*
All responses exclude “Don’t know” and skipped questions.
<sup>a</sup>Southern: Alabama, Arkansas, Delaware, DC, Florida, Georgia, Kentucky, Louisiana, Maryland, Mississippi, North Carolina, Oklahoma, South Carolina, Tennessee, Texas, Virginia, West Virginia
Northeastern: Connecticut, Maine, Massachusetts, New Hampshire, New Jersey, New York, Pennsylvania, Rhode Island, Vermont
Western: Alaska, Arizona, California, Colorado, Hawaii, Idaho, Montana, Nevada, New Mexico, Oregon, Utah, Washington, Wyoming
Midwestern: Indiana, Illinois, Iowa, Kansas, Michigan, Minnesota, Missouri, Nebraska, North Dakota, Ohio, South Dakota, Wisconsin

### COVID-19 Safety

Responses about COVID-19 safety measures and access to basic needs are reported for proxies and FII in aggregate (Table 2). Most participants (93.6%) reported experiencing lockdown procedures, of which 86.6% reported lockdowns >20 hours/day, and 48.8% reported a duration of >3 months. Over 92% of respondents indicated inability to always keep the CDC-recommended six-foot distance from others. Sixteen percent of respondents reported individual housing, while 53.7% reported housing with one other person, and 30% with two or more people. While 81.6% reported being provided PPE, (primarily face masks), 85.3% reported inconsistent use of PPE by staff. Over one-third of respondents (35.5%) reported that individuals who experienced COVID-19 symptoms were returned to their cells regardless of whether their cellmate also experienced symptoms. In addition, 53.5% reported that individuals were unable to take their belongings when being moved due to COVID-19. Lastly, nearly one-third (32.0%) of respondents reported that no information about COVID-19 was shared with individuals during incarceration. Among those who received information, the number of facility-wide COVID-19 cases was the most reported (32.8%).

**Table 2.**
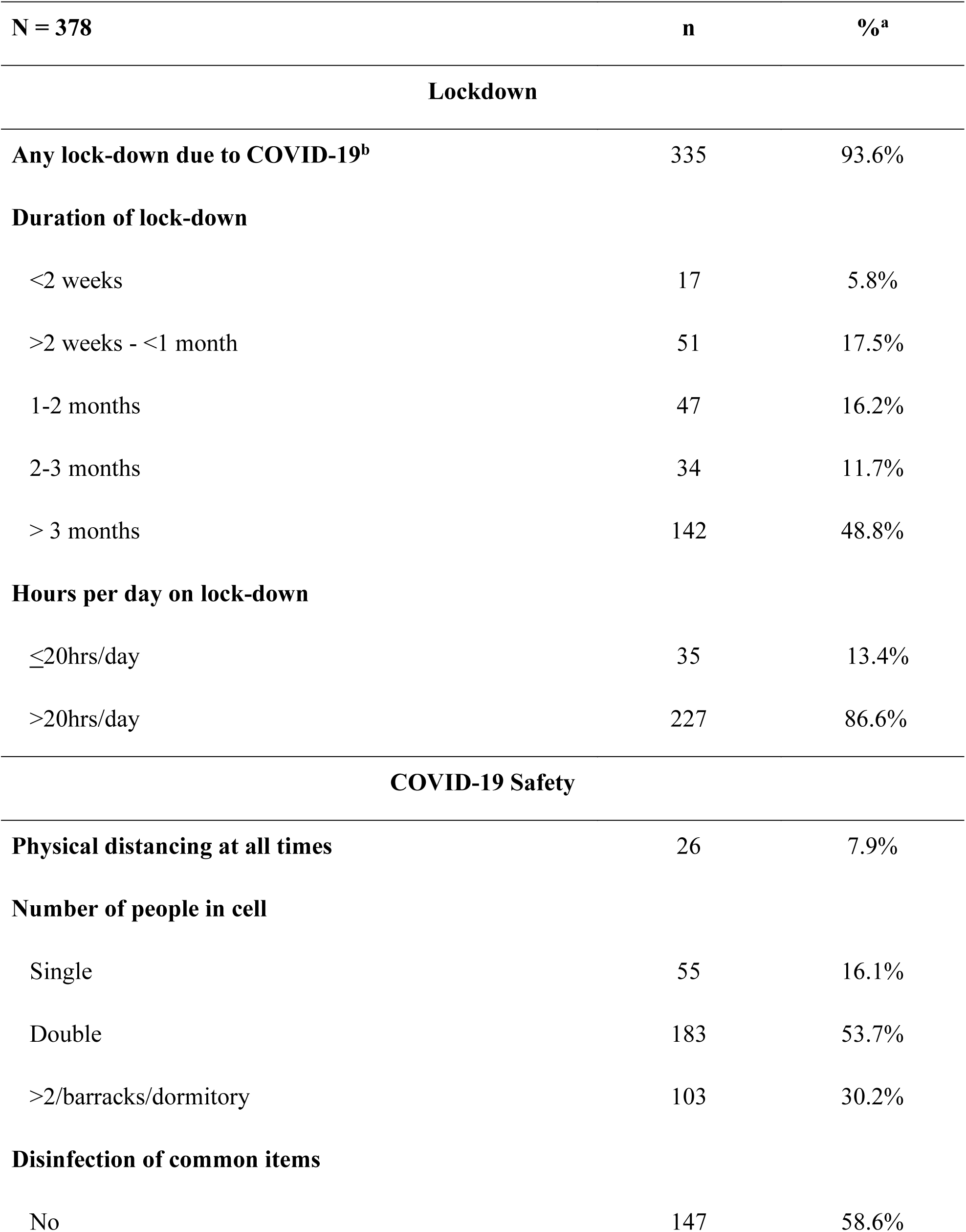
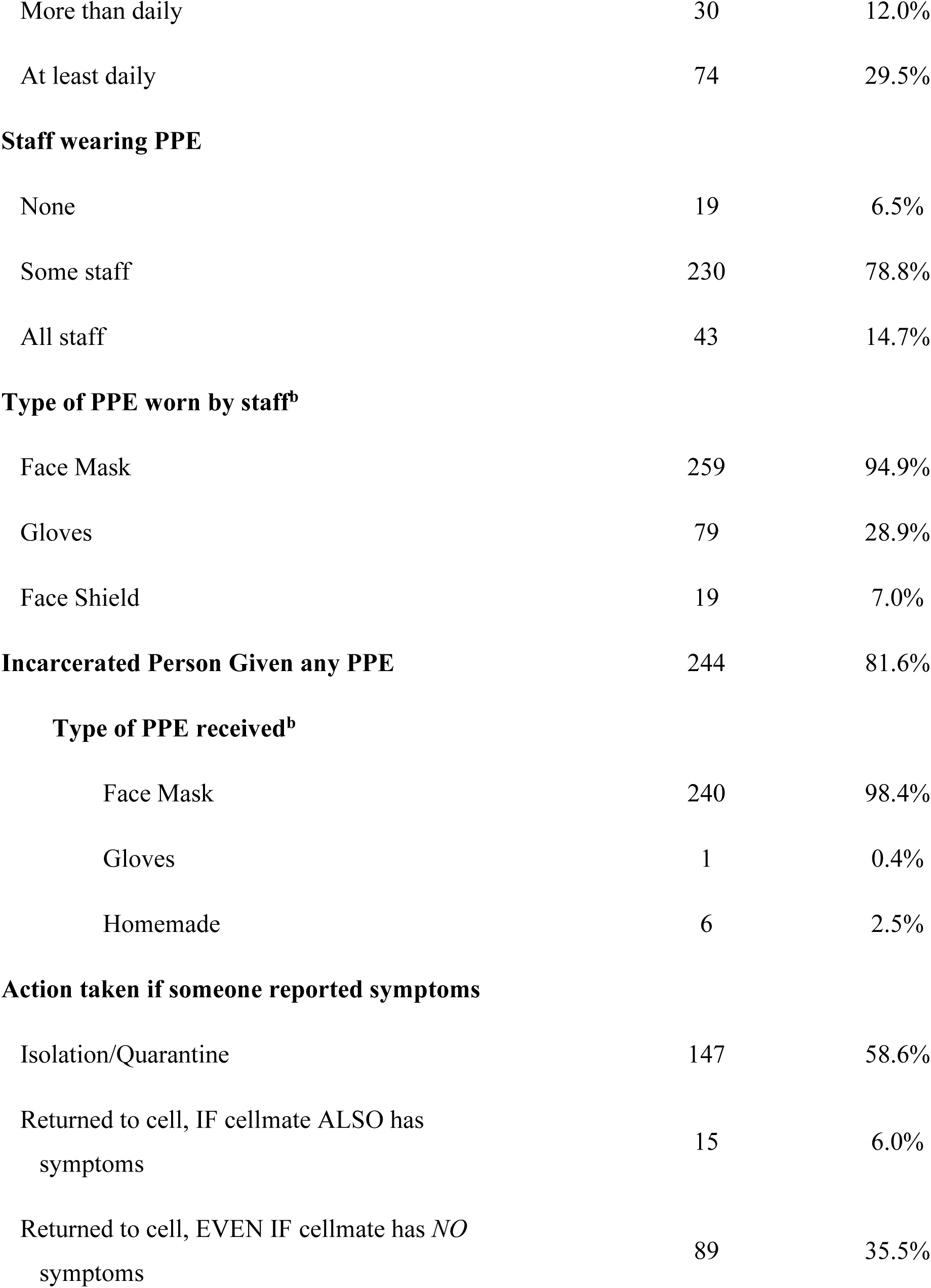
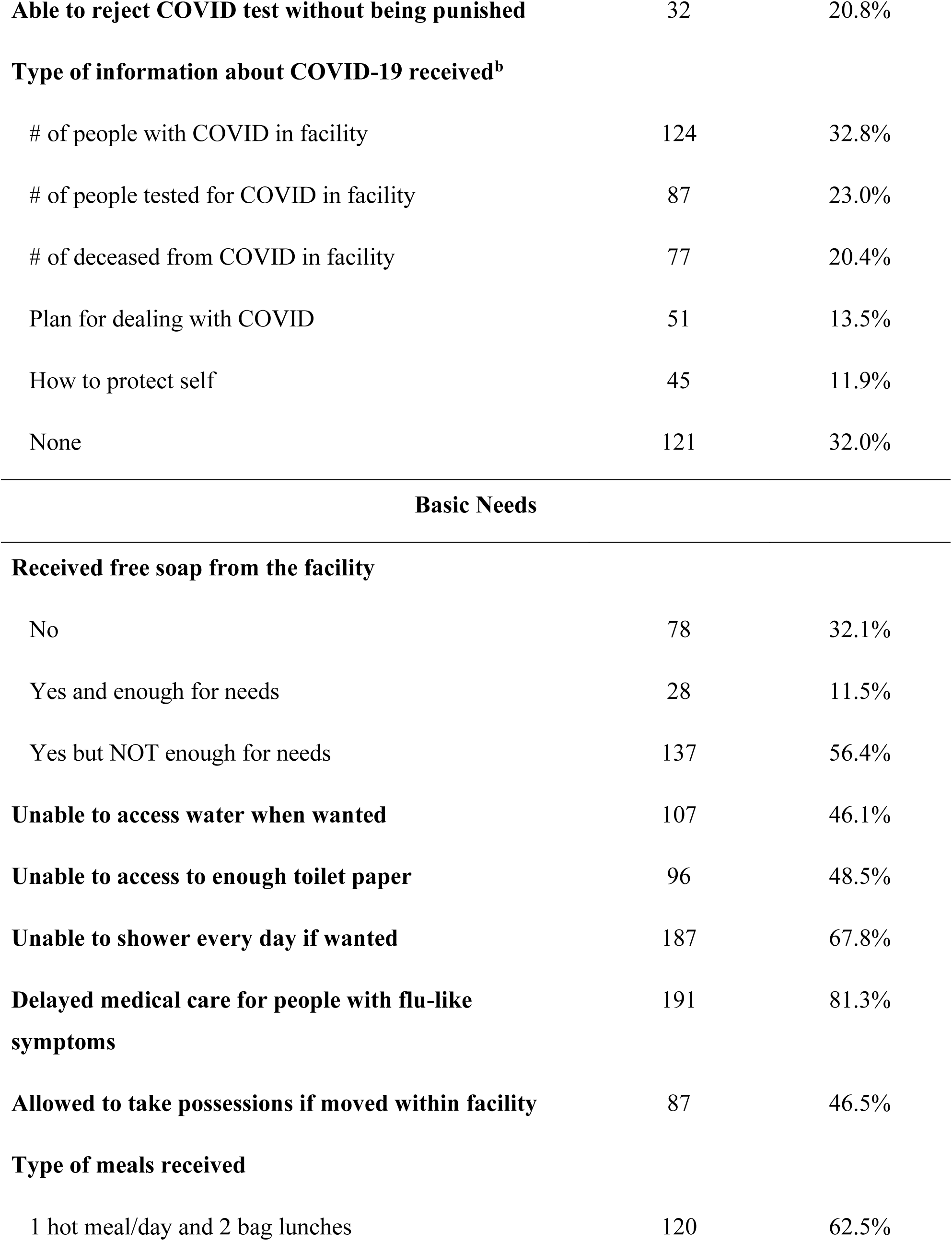
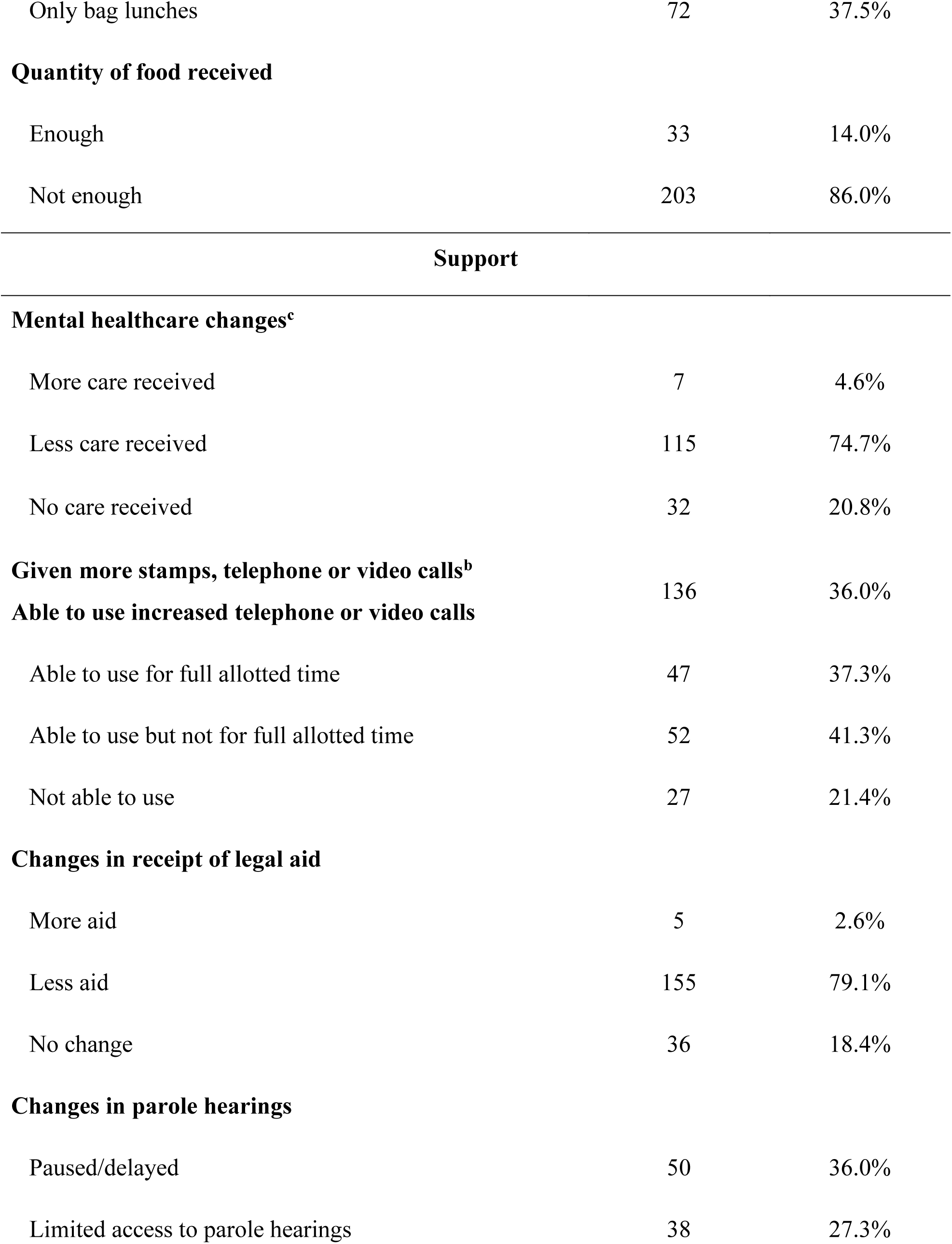
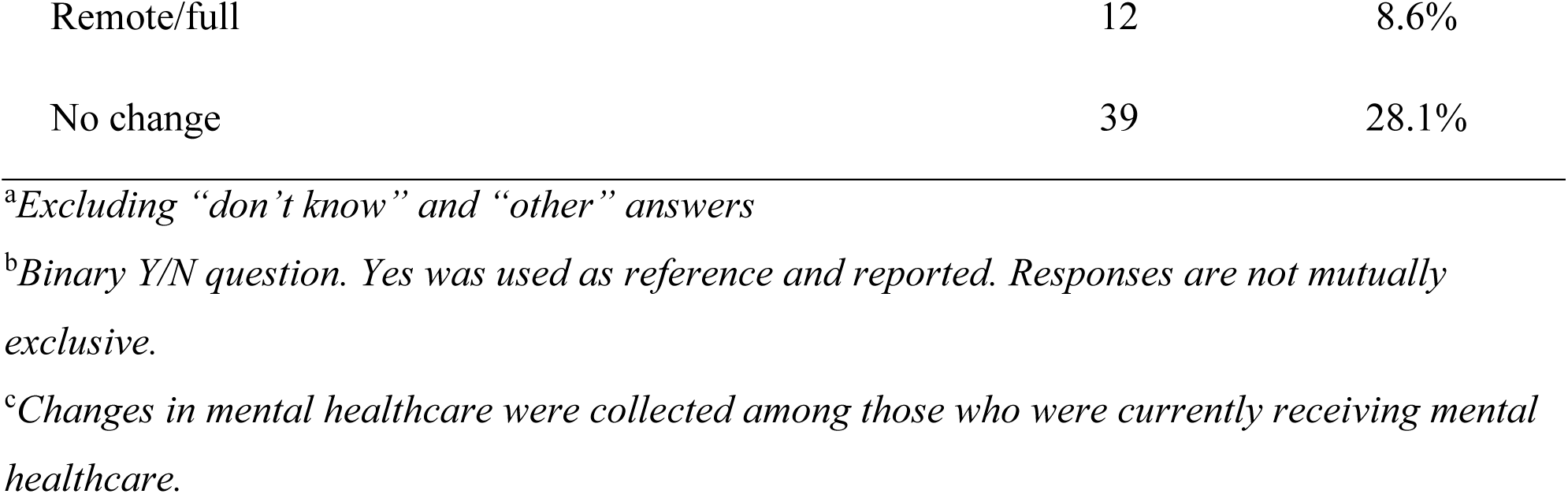
Distribution of Reported Conditions of Confinement.

| **N = 378** | **n** | **%<sup>a</sup>** |
| --- | --- | --- |
| **Lockdown** |  |  |
| **Any lock-down due to COVID-19<sup>b</sup>** | 335 | 93.6% |
| **Duration of lock-down** |  |  |
| <2 weeks | 17 | 5.8% |
| >2 weeks - <1 month | 51 | 17.5% |
| 1-2 months | 47 | 16.2% |
| 2-3 months | 34 | 11.7% |
| > 3 months | 142 | 48.8% |
| **Hours per day on lock-down** |  |  |
| ≤20hrs/day | 35 | 13.4% |
| >20hrs/day | 227 | 86.6% |
| **COVID-19 Safety** |  |  |
| **Physical distancing at all times** | 26 | 7.9% |
| **Number of people in cell** |  |  |
| Single | 55 | 16.1% |
| Double | 183 | 53.7% |
| >2/barracks/dormitory | 103 | 30.2% |
| **Disinfection of common items** |  |  |
| No | 147 | 58.6% |
| More than daily | 30 | 12.0% |
| At least daily | 74 | 29.5% |
| **Staff wearing PPE** |  |  |
| None | 19 | 6.5% |
| Some staff | 230 | 78.8% |
| All staff | 43 | 14.7% |
| **Type of PPE worn by staff<sup>b</sup>** |  |  |
| Face Mask | 259 | 94.9% |
| Gloves | 79 | 28.9% |
| Face Shield | 19 | 7.0% |
| **Incarcerated Person Given any PPE** | 244 | 81.6% |
| **Type of PPE received<sup>b</sup>** |  |  |
| Face Mask | 240 | 98.4% |
| Gloves | 1 | 0.4% |
| Homemade | 6 | 2.5% |
| **Action taken if someone reported symptoms** |  |  |
| Isolation/Quarantine | 147 | 58.6% |
| Returned to cell, IF cellmate ALSO has symptoms | 15 | 6.0% |
| Returned to cell, EVEN IF cellmate has *NO* symptoms | 89 | 35.5% |
| **Able to reject COVID test without being punished** | 32 | 20.8% |
| **Type of information about COVID-19 received<sup>b</sup>** |  |  |
| # of people with COVID in facility | 124 | 32.8% |
| # of people tested for COVID in facility | 87 | 23.0% |
| # of deceased from COVID in facility | 77 | 20.4% |
| Plan for dealing with COVID | 51 | 13.5% |
| How to protect self | 45 | 11.9% |
| None | 121 | 32.0% |
| **Received free soap from the facility** |  |  |
| No | 78 | 32.1% |
| Yes and enough for needs | 28 | 11.5% |
| Yes but NOT enough for needs | 137 | 56.4% |
| **Unable to access water when wanted** | 107 | 46.1% |
| **Unable to access to enough toilet paper** | 96 | 48.5% |
| **Unable to shower every day if wanted** | 187 | 67.8% |
| **Delayed medical care for people with flu-like symptoms** | 191 | 81.3% |
| **Allowed to take possessions if moved within facility** | 87 | 46.5% |
| **Type of meals received** |  |  |
| 1 hot meal/day and 2 bag lunches | 120 | 62.5% |
| Only bag lunches | 72 | 37.5% |
| **Quantity of food received** |  |  |
| Enough | 33 | 14.0% |
| Not enough | 203 | 86.0% |
| **Support** |  |  |
| **Mental healthcare changes<sup>c</sup>** |  |  |
| More care received | 7 | 4.6% |
| Less care received | 115 | 74.7% |
| No care received | 32 | 20.8% |
| **Given more stamps, telephone or video calls<sup>b</sup>** | 136 | 36.0% |
| **Able to use increased telephone or video calls** |  |  |
| Able to use for full allotted time | 47 | 37.3% |
| Able to use but not for full allotted time | 52 | 41.3% |
| Not able to use | 27 | 21.4% |
| **Changes in receipt of legal aid** |  |  |
| More aid | 5 | 2.6% |
| Less aid | 155 | 79.1% |
| No change | 36 | 18.4% |
| **Changes in parole hearings** |  |  |
| Paused/delayed | 50 | 36.0% |
| Limited access to parole hearings | 38 | 27.3% |
| Remote/full | 12 | 8.6% |
| No change | 39 | 28.1% |

<sup>a</sup>*Excluding “don’t know” and “other” answers*
<sup>b</sup>*Binary Y/N question. Yes was used as reference and reported. Responses are not mutually exclusive.*
<sup>c</sup>*Changes in mental healthcare were collected among those who were currently receiving mental healthcare.*

### Basic Needs, Medical Care, and Support

Overall, respondents indicated that individuals had inadequate access to basic necessities including personal hygiene items (Table 2). Although 67.9% received free soap, only 11.5% reported it was enough for their needs. Forty-six percent indicated insufficient access to water, toilet paper (48.5%) or daily showers (67.8%). In addition, 86.0% of respondents indicated insufficient food supply.

Medical care and support resources were also affected by the pandemic response (Table 2). A delay beyond normal delivery of medical care for flu-like symptoms was reported by 81.3% of respondents. Similarly, of those receiving mental healthcare before the pandemic, 74.7% reported a decrease access. Access to legal services were also reduced; 79.1% reported reductions in access to legal aid, and 36.0% indicated a delay of parole hearings.

### Questions only answered by Formerly Incarcerated Respondents

Questions about PPE replacement, worry and added stress, and receipt of financial support were asked only to formerly incarcerated respondents (n=21; Table 3). In total, 81.8% indicated new PPE was provided less than once per week. In addition, 87.5% reported that they worried about contracting COVID-19 “most of the time” and experienced “a lot of added stress” (93.8%). Most FII respondents (62.5%) received less financial support during the pandemic and only 16.7% reported that they received facility-level COVID-19 updates. Nearly two-thirds (64.3%) of FII expressed interest in getting a COVID-19 vaccine if available.

**Table 3.**
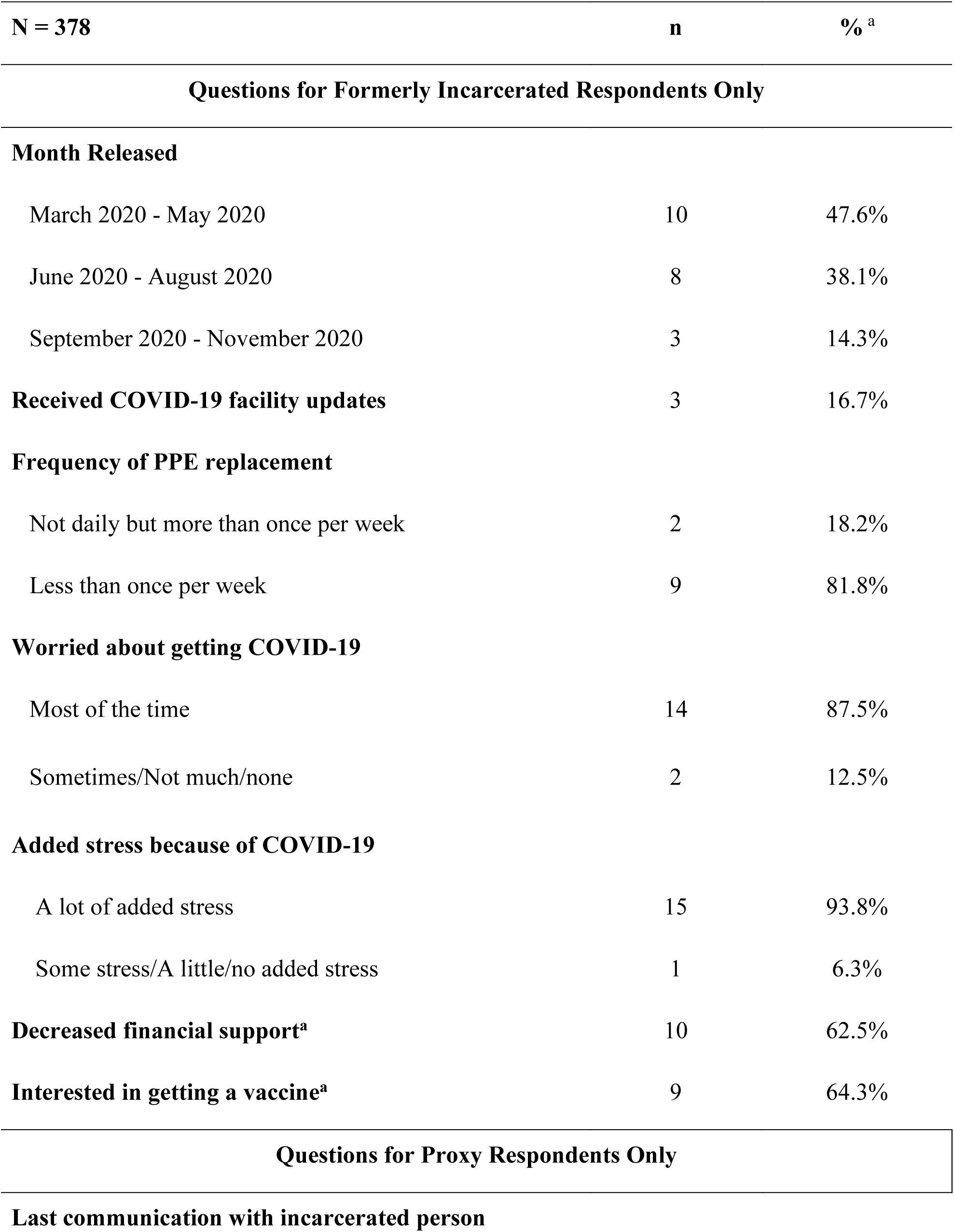
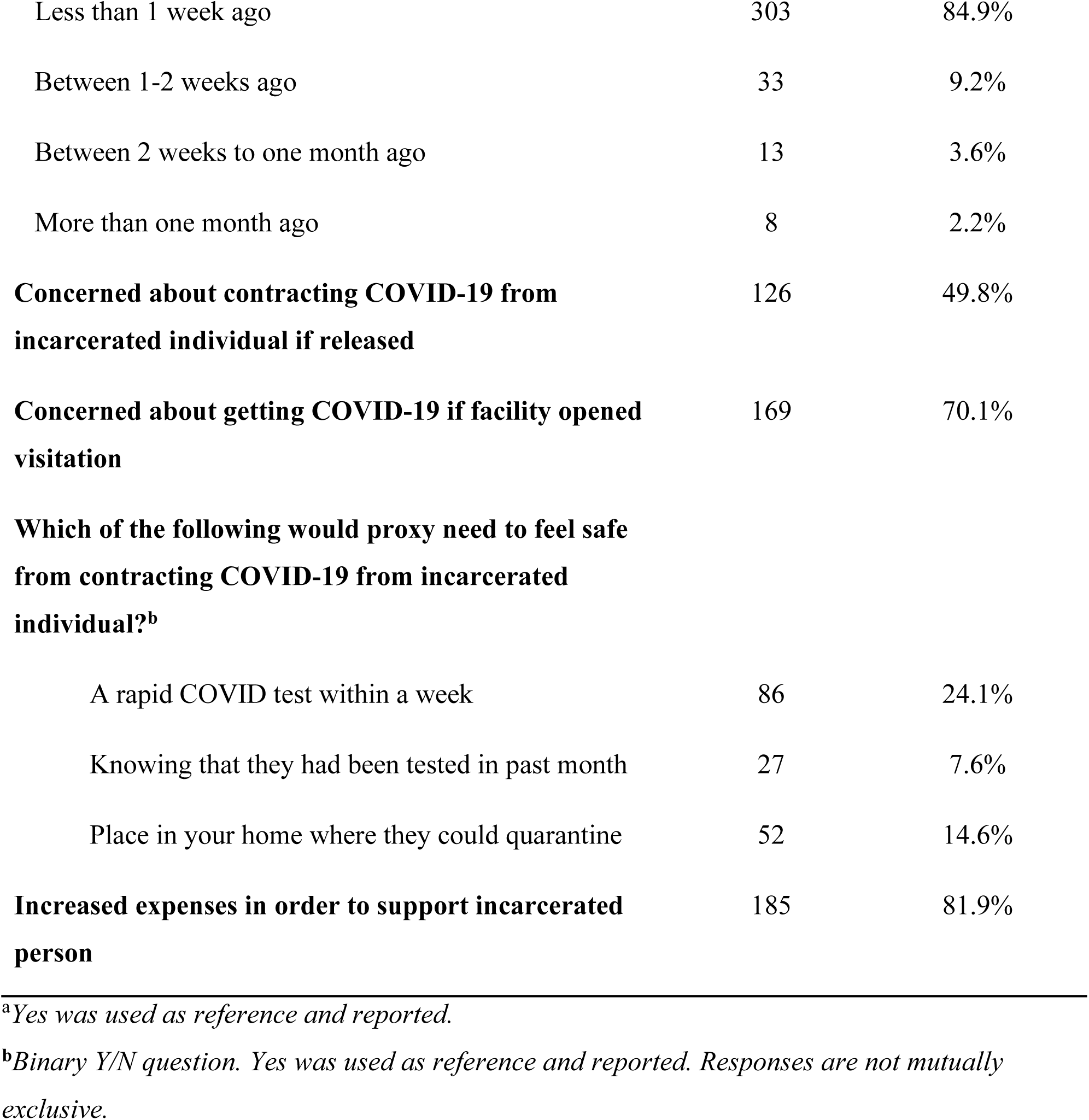
Questions Specifically for Formerly Incarcerated or Proxy Respondents.

| **N = 378** | **n** | **%<sup>a</sup>** |
| --- | --- | --- |
| **Questions for Formerly Incarcerated Respondents Only** |  |  |
| **Month Released** |  |  |
| March 2020 - May 2020 | 10 | 47.6% |
| June 2020 - August 2020 | 8 | 38.1% |
| September 2020 - November 2020 | 3 | 14.3% |
| **Received COVID-19 facility updates** | 3 | 16.7% |
| **Frequency of PPE replacement** |  |  |
| Not daily but more than once per week | 2 | 18.2% |
| Less than once per week | 9 | 81.8% |
| **Worried about getting COVID-19** |  |  |
| Most of the time | 14 | 87.5% |
| Sometimes/Not much/none | 2 | 12.5% |
| **Added stress because of COVID-19** |  |  |
| A lot of added stress | 15 | 93.8% |
| Some stress/A little/no added stress | 1 | 6.3% |
| **Decreased financial support<sup>a</sup>** | 10 | 62.5% |
| **Interested in getting a vaccine<sup>a</sup>** | 9 | 64.3% |
| **Questions for Proxy Respondents Only** |  |  |
| **Last communication with incarcerated person** |  |  |
| Less than 1 week ago | 303 | 84.9% |
| Between 1-2 weeks ago | 33 | 9.2% |
| Between 2 weeks to one month ago | 13 | 3.6% |
| More than one month ago | 8 | 2.2% |
| **Concerned about contracting COVID-19 from incarcerated individual if released** | 126 | 49.8% |
| **Concerned about getting COVID-19 if facility opened visitation** | 169 | 70.1% |
| **Which of the following would proxy need to feel safe from contracting COVID-19 from incarcerated individual?<sup>b</sup>** |  |  |
| A rapid COVID test within a week | 86 | 24.1% |
| Knowing that they had been tested in past month | 27 | 7.6% |
| Place in your home where they could quarantine | 52 | 14.6% |
| **Increased expenses in order to support incarcerated person** | 185 | 81.9% |

<sup>a</sup>*Yes was used as reference and reported.*
<sup>b</sup>*Binary Y/N question. Yes was used as reference and reported. Responses are not mutually exclusive.*

**Table 4.** Internal Validity Results.

| **Question** | **Proxy Respondents** | **Formerly Incarcerated Respondents** | **Two-sided Fisher's exact test p-value** |
| --- | --- | --- | --- |
| Facility on any type of lockdown | 94.4% (320) | 79.0% (15) | **0.03** |
| Hours spent in lockdown (>20 hours/day) | 86.2% (213) | 93.3% (14) | 0.70 |
| Lockdown time period exceeding 3 months | 49.6% (137) | 33.3% (5) | 0.29 |
| Ability to maintain 6-foot distance at all times | 7.8% (24) | 10.0% (2) | 0.67 |
| Routine disinfection of commonly used surfaces | 40.6% (95) | 52.9% (9) | 0.32 |
| Adequate access to toilet paper | 50.3% (90) | 63.2% (12) | 0.34 |
| Ability to shower every day, if wanted | 30.0% (77) | 63.2% (12) | **0.01** |
| Adequate access to water | 53.5% (114) | 57.9% (11) | 0.81 |
| Enough free soap for needs | 10.3% (23) | 26.3% (5) | 0.05 |
| Isolated/Quarantined from cellmates if experiencing COVID-19 symptoms | 59.2% (142) | 45.5% (5) | 0.37 |
| Longer waits than normal to access medical care | 81.2% (177) | 82.4% (14) | 1.00 |
| Increased access to mental healthcare | 4.1% (6) | 11.1% (1) | 0.35 |

*All responses are dichotomized; values represent responses to “yes.”*

### Questions only answered by Proxy Respondents

Questions about recency of communication with incarcerated individuals, concerns about contracting COVID-19 from FII when released or through visitation, and increased expenses to support incarcerated individuals were posed specifically to proxies. Most had communicated with the individual within the past week (85%). A high proportion (70.1%) of proxies reported concerns about contracting the virus when facilities reopened for visitation or from the incarcerated individual if released (49.8%). Nearly a quarter of respondents (24.1%) reported needing a rapid COVID-19 test within a week to feel safe if the incarcerated individual were released tomorrow. Additionally, 81.9% reported increased expenses to support the individual.

Since respondents were allowed to skip questions, missing data ranged from 4.7% for facility lockdown to 59% for access to mental healthcare.

### Internal Validity

The distribution of responses between proxies and FII were compared for key metrics and were found to be consistent (p>0.05) for all but two questions. First, proxies were significantly more likely to report a facility being on lockdown (94.4%) than FII (79.0%, p=0.03). Additionally, fewer proxies (30.0%) reported that incarcerated individuals had access to daily showers compared to 63.2% of FII (p=0.01).

## DISCUSSION

To our knowledge, this is the first study designed in partnership with FII to collect data on confinement conditions among individuals incarcerated across all four regions of the U.S. Our findings suggest that at least in some facilities, individuals were unable to always physically distance from others, despite extensive periods of lockdown, and lacked access to basic needs (i.e., soap, water, toilet paper, and showers). Respondents also indicated limited access to healthcare and legal aid.

One other quantitative study has reported confinement conditions based on 327 individuals incarcerated in three U.S. states.^19^ The COVID-19 Questionnaire for Correctional Populations (CQCP) survey was used to collect data on mitigation strategies observed by staff and incarcerated individuals. Our findings were similar to those reported by the CQCP study for some conditions, such as provision of soap (INSIDE: 67.9%; CQCP: 70.6%). However, we observed discrepancies for questions regarding physical distancing (INSIDE: 7.9%; CQCP: 66.4%), which may be due to wording differences. While the CQCP asked if participants “Stayed 6 feet away from others if possible,” we explicitly asked, “Were you allowed to keep your distance from others by staying 6 feet away at all times?”. Our study complements and expands this previous work by collecting nationwide data, reflecting a longer time-period (July 2020-March 2021 versus April 2020-November 2020) and includes questions on lockdown, basic needs, and mental healthcare access.

Our findings also paralleled several studies from the gray literature. The Essie Justice Group surveyed 729 people who were in contact with an incarcerated individual.^22^ Overall, 62% reported that their incarcerated loved ones feared losing their lives, similar to 87.5% of FII from INSIDE who worried about getting COVID-19 “most of the time”. Similarly, a Physicians for Human Rights survey among 50 individuals formerly detained by ICE, reported that 80% were not able to maintain a 6-foot distance from others while eating, consistent with the 92% of our respondents who reported an inability to distance ≥6-feet at all times.^15^

Our data suggest that implementation of COVID-19 mitigation strategies may not meet CDC-outlined practices and may lead to physical and psychological distress. For example, numerous studies have shown solitary confinement’s association with psychological harm^23-25^ including anxiety, depression, aggression, self-harm, and increased mortality post-release.^23^ Yet, lockdowns which result in extended confinement and isolation have been prioritized as a strategy to reduce transmission. One INSIDE FII reported,

> *“Being locked in a cell for 23 hours and 40 minutes a day for weeks at a time (whenever there was a report of a positive case among staff/incarcerated citizens was extremely stressful. I often considered hurting other prisoners or the guards simply because I was angry. I am not a violent person. I was incarcerated for a drug offense.”*

In addition, in the general population, post-pandemic delayed or restricted care have likely contributed to the 18% increase in mortality (2020 versus 2019).^26^ Specifically, mortality increases in diabetes (15%), unintentional injury (11%), Alzheimer’s (10%), and heart disease (5%) are suggestive of collateral pandemic consequences.^26^ In our study, prolonged time to receive medical care (81.3%) and reduced access to mental health care (74.7%) may be associated with increased risk of death or morbidity either from COVID-19 directly or chronic conditions. One proxy respondent reported,

> *“My loved one recently got infected with COVID-19 and is also suffering for underlying medical conditions (chronic Hepatitis B and asthma). He didn’t receive any medical treatments other than having his inhaler with him… The only “treatment” that was given to some individuals were over-the-counter medications such as Tylenol to reduce some flu-like symptoms. My loved one is still suffering from some lingering symptoms such as wheezing and coughing.”*

Our findings support recommendations from public health professionals and advocacy groups to reform crisis responses strategies by centering (1) population decompression, (2) improvement of living conditions through external oversight, and (3) incorporation of input from incarcerated people and their families.^27,28^ Importantly, the American Public Health Association has called for wide-spread population decompression due to the well-documented harms associated with incarceration which have only been exacerbated by the pandemic.^27^ Their comprehensive recommendations include use of diversion programs, sentencing, misdemeanor and bail reform. Additional tools for population decompression include expansion of compassionate release for those with chronic medical conditions and the elderly, home confinement programs, and expedited release of those nearing their release date.^29^

Our data also demonstrated some facilities’ failure to provide basic needs, medical and mental healthcare, and implement safety measures to mitigate infectious disease transmission. However, there is no central external oversight body for US prisons and jails, resulting in limited accountability, inconsistent crisis response, and a lack of standardized protections.^30^ In order to minimize harm to incarcerated people, transparent external oversight is necessary to ensure humane adherence to CDC guidelines. Lastly, our study provides a model for how directly impacted people and their loved ones can provide critical evidence regarding implementation of crisis response strategies, while highlighting the need for anonymous collection of firsthand experiences to foster transparency and ensure compliance.

To our knowledge, this study is the only study to quantitatively document confinement conditions during the pandemic with data from all four U.S. regions and across a variety of facility types. Importantly, this study was conducted in partnership with a CAB and driven by community-identified needs. Lastly, we demonstrate the feasibility and validity of gathering data on the lived experiences of incarcerated individuals via proxies.

We acknowledge that generalizability of our findings may be limited using a convenience sample. However, we aimed to increase geographic and carceral system diversity and recognized that obtaining required approvals from each independent facility to directly survey incarcerated individuals would hinder the collection of time-sensitive data. Despite this limitations, general comparisons with other studies and anecdotal reports support the patterns we observed. In addition, generalizability of our sample by race and ethnicity is uncertain due to missing data for these variables. Although our sample is likely to overrepresent conditions in female facilities given our CAB member’s networks; there is no evidence to suggest that conditions would be improved for incarcerated males or individuals of color. Furthermore, our novel data collection methods were developed early in the pandemic based on public health priorities at that time which have changed with respect to sanitation procedures over the course of the pandemic.

Furthermore, data from proxies may not be optimally reported for all variables due to the proportion of “I don’t know” or “Skip” responses to some questions, such as mental healthcare access. Lastly, two of our variables (facility on lockdown and ability to shower when wanted) demonstrated low internal validity; significantly more FII reported ability to shower daily if wanted, compared to proxies, while more proxies reported lockdown conditions compared to FII. However, 15.8% of FII who answered the questions regarding showering and lockdown were released before May 2020, prior to widespread response in all states. Additionally, all were incarcerated in Florida, which may reflect variability in different states’ responses. However, we observed high internal validity for other variables, suggesting that proxy reports largely matched those of incarcerated individuals.

## HEALTH EQUITY IMPLICATIONS

Our data demonstrate that at least some incarcerated people experienced inadequate implementation of safety protocols, difficulty accessing basic needs, and were subjected to long periods of restricted confinement. Importantly, our data highlight the need to prioritize the lived experiences of incarcerated individuals in policy design and oversight, to ensure appropriate and humane policies and implementation. Our data were collected through robust community partnerships, and we urge practitioners to engage with community organizations to develop meaningful research questions, leverage community strengths, and shape policy.

## Supporting information

SurveyTool

STROBEChecklist

## Data Availability

All data produced in the present study are available upon reasonable request to the authors

## Acknowledgements

Dr. Jiménez is supported by the Brigham and Women’s Hospital Richard Nesson Fellowship. The authors would like to thank Massachusetts State Senator Rebecca Rausch, communications director Evan Berry, and intern Sandy Mabee for their contributions to this study.

