## Supplementary material for "Conditions of Confinement in U.S. Carceral Facilities during COVID-19: Individuals Speak: Incarcerated during the COVID-19 Epidemic (INSIDE)": SurveyTool

**Survey instrument**

### Role of respondent and Facility

1 **Please choose which statement describes you: (X)**

- I am filling out the survey based on what I know from recently talking to someone who is incarcerated now
    - i. *(FF):* These questions will be about the experience of the person you know who is incarcerated now. These questions ask about their experiences starting from March 1, 2020. You can skip any questions if you do not know the answer. Please do not share any personal information like name, date of birth, or other identifying information. For example, please do not report your own name or the name or other personal information of the person you know who is incarcerated.
    - ii. **When was the last time you communicated to this person**? (X)
        - Less than 1 week ago
        - More than a week but less than 2 weeks ago
        - More than 2 weeks but within the past month
        - More than one month ago {End survey}

- I am filling out the survey based on my own experience
    - i. *(FI)*: **Were you released from a correctional or detention facility after March 1, 2020?** (X)
        - **Yes:** These questions are about your experience of incarceration as early as March 1, 2020 up until the time you were released. If you were incarcerated for any length of time between March 1, 2020 and today you can take this survey. You can skip any questions if you do not know the answer. Please do not report any personal information like your name, date of birth, or other identifying information. For example, please do not report your own name or date of birth or details related to the reason for your incarceration.
            - **What month were you released**? (X)
                - March 2020
                - April 2020
                - May 2020
                - June 2020
                - July 2020
                - August 2020
        - **No:** We value your experience and feedback, but these questions are only about conditions in correctional facilities since the COVID-19 pandemic started. If you can share information based on talking to someone who is incarcerated now, please go back to question 1 and choose "I am filling out the survey based on what I know from talking to someone who is currently incarcerated." {End Survey}

2 **FI: What state were you incarcerated in:** [Drop down menu] (x)
**FF: What state is the person incarcerated in:** [Drop down menu] (x)

3 **FI: What type of facility were you incarcerated in after March 1, 2020: (X)**
**FF: What type of facility is the person currently incarcerated in:**

- Jail
- State Prison
- Federal Prison
- ICE Detention Center
- Other: [text]
- Skip

| **The following questions are about being allowed to keep physical distance from others.** | |
|---|---|
| 4 | **FI: Was the <u>facility</u> on any type of lock-down because of COVID-19?** (Lock-down is any type of restricted movement out of a cell, barracks or dorm that is applied to the whole facility) **(X)**<br>**FF: Is their <u>facility</u> on any type of lock-down because of COVID-19?** (Lock-down is any type of restricted movement out of a cell, barracks or dorm that is applied to the whole facility) **(x)**<br>● Yes<br>● No<br>● Other: [text]<br>● I don't know<br>● Skip |
| 5 | **FI: How long were you on lock-down? (X)**<br>**FF: How long have they been on lock-down? (X)**<br>● Less than two weeks<br>● More than two weeks but less than a month<br>● One to two months<br>● Two to three months<br>● Three months or more<br>● None<br>● Skip<br>● I don't know |
| 5a | **FI: How many hours per day were you on lock-down?** (Please do not leave this blank. Choose 'Skip' if you are unsure.) [numeric drop down] (x)<br>**FF: How many hours per day are they on lock-down?** (Please do not leave this blank. Choose 'Skip' if you are unsure.) [numeric drop down] (X) |
| 6 | **Facilities are stating they are taking steps to protect incarcerated people from getting COVID-19. We would like to know, (x)**<br>**FI: Were you allowed to keep your distance from others by staying 6 feet away from others <u>at all times</u>?**<br>**FF: In the past month, is the person you know allowed to keep their distance from others by staying 6 feet away <u>at all times</u>?**<br>● Yes<br>● No<br>● I don't know<br>● Skip |
| 7 | **FI: How many people were in your cell, including yourself? (X)**<br>**FF: In the past two weeks, how many people, including the person, were in their cell? (X)**<br>● 1 (single cell)<br>● 2 (double cell)<br>● More than a two-person cell or barracks or dorm<br>● Other [text]<br>● I don't know<br>● Skip |

| | |
|---|---|
| **The following questions are about the ability to live in a clean environment.** | |
| 8 | **FI: Did you get free soap from the facility? Please do not include soap provided or shared by other incarcerated individuals. (X)**<br>**FF: In the past month, has the person gotten free soap from the facility? Please do not include soap provided or shared by other incarcerated individuals. (X)**<br>• Yes, enough for my needs<br>• Yes, but NOT enough for my needs<br>• No<br>• I don't know<br>• Skip |
| 9 | **FI: Were you able to get water when you wanted? (X)**<br>**FF: In the past month, are they able to get water when they want? (X)**<br>• Yes<br>• No<br>• I don't know<br>• Skip |
| 10 | **FI: Were things that were commonly touched or used being disinfected? For example, doors, phones, microwaves.** (X)<br>**FF: In the past month, have things that are commonly touched or used been disinfected? For example, doors, phones, microwaves.** (X)<br>• Yes, multiple times a day<br>• Yes, one time a day<br>• No<br>• I don't know<br>• Skip |
| 11 | **FI: Were you able to get enough toilet paper? (X)**<br>**FF: In the past month, have they been able to get enough toilet paper? (X)**<br>• Yes<br>• No<br>• I don't know<br>• Skip |
| 12 | **FI: Were you able to shower every day, if you wanted to? (X)**<br>**FF: In the past month, could they shower every day, if they wanted to? (X)**<br>• Yes<br>• No<br>• I don't know<br>• Skip |
| **The following questions are about special equipment needed to protect yourself from COVID-19.** | |
| 3 | **FI: Were staff wearing any type of protection from COVID-19? Examples are face masks, face shields, gloves, protective clothing. We will ask about which types in the next question. (X)**<br>**FF: In the past month, have staff been wearing any type of protection from COVID-19? Examples are face masks, face shields, gloves, protective clothing. We will ask about which types in the next question. (X)**<br>• Yes, all staff<br>• Some staff<br>• No, none of the staff<br>• I don't know<br>• Skip |

<table>
<tr><td>13a</td><td>FI: What types of protective gear did they wear? Click all that apply. (X)<br>FF: What types of protective gear are they wearing? Click all that apply. (X)<br>● Face masks<br>● Gloves<br>● Face shield (clear plastic face covering)<br>● Other: [TEXT]<br>● Skip</td></tr>
<tr><td>14</td><td>FI: Were you given anything to wear to protect yourself from COVID-19? (X)<br>FF: Was the person you know given anything to wear to protect them from COVID-19? (X)<br>● Yes<br>● No<br>● I don’t know<br>● Skip</td></tr>
<tr><td>14a</td><td>FI: What type of protective gear were you given? <u>Please check all that apply (X)</u><br>FF: What type of protective gear were they given? <u>Please check all that apply(X)</u><br>● Face masks<br>● Gloves<br>● Not given something but allowed to make one<br>● Other:<br>● Skip</td></tr>
<tr><td>14b</td><td>FI: How often were you given new protective gear? (X)<br>● Daily<br>● Not daily, but at least once a week<br>● Less than once a week<br>● I don’t know<br>● Skip</td></tr>
<tr><td colspan="2">The following questions are about medical care.</td></tr>
<tr><td>15</td><td>FI: What happened if someone reported symptoms [such as cough, fever, sore throat, loss of smell or taste, soreness]? (X)<br>FF: In the last month, what happens if someone reports symptoms [such as cough, fever, sore throat, loss of smell or taste, soreness]? (X)<br>● The person is separated (placed in isolation or quarantine)<br>● The person is returned to their regular housing only if cellmate(s) ALSO have symptoms<br>● The person is returned to their regular housing even if cellmate(s) DO NOT have symptoms<br>● I don’t know<br>● Other: [text]<br>● Skip</td></tr>
<tr><td>16</td><td>FI: Did it take longer than normal for people to get medical care if they had flu-like symptoms [such as cough, fever, sore throat, loss of smell or taste, soreness]? (X)<br>FF: Does it take longer than normal for people to get medical care if they have flu-like symptoms [such as cough, fever, sore throat, loss of smell or taste, soreness]? (X)<br>● Yes<br>● No<br>● I don’t know<br>● Skip</td></tr>
</table>

| | |
|---|---|
| 17 | **FI: If you received mental health care, was the type of treatment changed because of COVID-19? (X)**<br>**FF: If the person you know receives mental health care, has the type of treatment changed because of COVID-19? (X)**<br>• Yes, I have gotten more care<br>• Yes, I have gotten less care<br>• No, care has not changed<br>• I don't know<br>• Does not apply<br>• Skip |
| 18 | **FI: If someone was moved because of COVID-19 were they allowed to take their possessions with them? (X)**<br>**FF: If someone is moved because of COVID-19 are they allowed to take their possessions with them? (X)**<br>• Yes<br>• No<br>• I don't know<br>• Skip |
| 19 | **FI: If someone wanted to be tested were they able to get tested? (X)**<br>**FF: If someone wants to be tested are they able to get tested? (X)**<br>• Yes<br>• No<br>• I don't know<br>• Skip |
| 20 | **FI: If someone did not want to be tested, could they say no without being punished? (X)**<br>**FF: If someone does not want to be tested, can they say no without being punished? (X)**<br>• Yes<br>• No<br>• I don't know<br>• Skip |
| **The following questions are about information you were given about COVID-19.** | |
| 21 | **FI: What type of information was shared with you by staff about COVID-19 where you were incarcerated? <u>Please check all that apply</u>**<br>**FF: What type of information was shared with you about COVID-19 by the facility where the person you know is incarcerated? <u>Please check all that apply</u>**<br>• The number of people with COVID-19 in the facility<br>• The number of people tested for COVID-19 in the facility<br>• The number of people who have died from COVID-19 in the facility<br>• The plan for dealing with COVID-19 in the facility<br>• FI: How to protect yourself / FF: How they can protect themselves<br>• None<br>• I don't know<br>• Skip |
| 21a | **FI: Did you get updates about COVID-19 in the facility? (X)**<br>• Yes<br>• No<br>• I don't know<br>• Skip |

| | |
|---|---|
| 22 | **FI: If a vaccine were available would you get it? (X)**<br>● Yes [Skip #21]<br>● No<br>● I don't know [Skip #21]<br>● Skip |
| 22a | **FI: If no, could you tell us why?** (To skip this question, please type: NA) **[text]** |
| **The following questions are about well-being.** | |
| 23 | **FI: Can you tell us what gave you strength through the day while you were incarcerated during this time?** (To skip this question, please type: NA) |
| 24 | **FI: How often did you find yourself worried about getting COVID-19?** (X)<br>● Most of the time<br>● Sometimes<br>● Not much or not at all<br>● I don't know<br>● Skip |
| 25 | **FI: How much added stress, tension, or difficulties did you feel because of COVID-19 while you were incarcerated? (X)**<br>● A lot of added stress<br>● Some stress<br>● A little or no added stress<br>● I don't know<br>● Skip |
| 26 | **FI: Which of the following statements best describes the type of meals you received while you were incarcerated during COVID-19? (X)**<br>**FF: Which of the following statements best describes the type of meals they have received in the past month? (X)**<br>● Getting at least one hot meal a day and two bag lunches<br>● Not getting any hot meals, only bag lunches<br>● I don't know<br>● Other: [text]<br>● Skip |
| 27 | **FI: Which of the following statements best describes the quantity of food received while you were incarcerated during COVID-19? (x)**<br>**FF: Which of the following statements best describes the quantity of food received in the past month? (x)**<br>● Getting enough food to meet needs<br>● NOT getting enough food to meet needs<br>● I don't know<br>● Other: [text]<br>● Skip |
| 28 | **FF: If your family member could get out tomorrow do you have any concerns about getting COVID-19 from them? (x)**<br>● Yes<br>● No [skip next question]<br>● I don't know<br>● Skip |

| | |
|---|---|
| 28a | **FF: What would you need to feel safe from getting COVID-19? Please check all that apply. (x)**<br>● A rapid covid test within a week<br>● Knowing that they had been tested in the past month<br>● Place in your home where they could quarantine<br>● Something else: [text]<br>● Skip |
| 29 | **FF: If the facility where the person you know is incarcerated opened visitation, would you have any concerns about getting COVID-19? (x)**<br>● Yes<br>● No<br>● I don't know<br>● Skip |
| 30 | **FI: If you received financial support from friends or family, did that decrease due to COVID-19? (X) FF: Have you had more expenses or bills than normal to help support the person you know who is incarcerated during this time? (X)**<br>● Yes<br>● No<br>● I don't know<br>● Skip |
| **The following questions are about the ability to stay in communication with family, friends or legal aid.** | |
| 31 | **Some facilities have stated that they have given people more calls or stamps to keep in touch with family and friends.**<br><br>**FI: Were you given more telephones calls, video calls or stamps for mail? <u>Check all that apply</u>.**<br>**FF: Have they been given more telephones calls, video calls or stamps for mail? <u>Check all that apply</u>.**<br>● More telephone calls<br>● More video calls<br>● More stamps {Skip to #29}<br>● None {Skip to #29}<br>● I don't know<br>● Skip |
| 31a | **FI: Were you able to use the increased telephone or video calls?**<br>**FF: Are they able to use the increased telephone or video calls?**<br>● Yes, for the full amount of time<br>● Yes, but calls are shorter than usual<br>● Not able to use extra calls<br>● I don't know<br>● Skip |
| 32 | **FI: Were there changes to your ability to get legal aid? (X)**<br>**FF: Have they seen changes in their ability to get legal aid? (X)**<br>● No changes<br>● More access to legal aid<br>● Less access to legal aid<br>● I don't know<br>● Skip |

| | |
|---|---|
| 33 | **FI: Were there changes to your parole hearings? (X)**<br>**FF: Have they seen changes in their parole hearings? (X)**<br>● Parole hearings paused or delayed<br>● Limited access to parole hearings<br>● Remote parole hearings with full access<br>● No changes<br>● I don't know<br>● Other: [text]<br>● Skip |
| 34 | **FI: If you were enrolled in programming to earn good time credits, have there been changes to being allowed to get those credits?**<br>● No option to get good time credit<br>● Some good time credit but not full credit<br>● Getting good time credits has not been changed<br>● I don't know<br>● I was not enrolled<br>● Skip |
| **We'd like to learn more about these experiences. Please write in anything you would like us to know.** *Please <u>do not</u> include any private information such as name, birthday, address, telephone number, booking number, "inmate ID" or social security number.* | |
| 35 | **FI: Can you tell us about some of the ways in which you tried to protect yourself from getting COVID-19?** (To skip this question, please type: NA)<br>**FF: Can you tell us about some of the ways that they try to protect themselves from getting COVID-19?** (To skip this question, please type: NA) |
| 36 | **FI: Have you heard about any incarcerated person who has died of COVID-19? What were the circumstances?** (To skip this question, please type: NA)<br>**FF: Have they heard about any incarcerated person who died of COVID-19? What were the circumstances?** (To skip this question, please type: NA) |
| 37 | **FI: How much do you trust information about COVID-19 that was shared with you by the facility?** (To skip this question, please type: NA)<br>**FF: How much do you trust information about COVID-19 that was shared with you by the facility?** (To skip this question, please type: NA) |
| 38 | **Is there anything else you would like to share with us?** (To skip this question, please type: NA) |
| **The next questions are about your background.** | |
| 39 | **FI: Do you think of yourself as**: (x)<br>**FF: Do they think of themselves as**: (x)<br>● Male<br>● Female<br>● Transgender man / trans man / female-to-male<br>● Transgender woman / trans woman / male-to-female<br>● Genderqueer / Gender nonconforming neither exclusively male nor female<br>● Additional gender category or other (please specify): [text]<br>● I don't know<br>● Skip |

| | |
|---|---|
| 40 | **FI: Are you Hispanic, Latino/a/x or of Spanish origin** (a person of Cuban, Mexican, Puerto Rican, South or Central American, or other Spanish culture or origin, regardless of race)? (x)<br>**FF: Are they Hispanic, Latino/a/x or of Spanish origin** (a person of Cuban, Mexican, Puerto Rican, South or Central American, or other Spanish culture or origin, regardless of race)? (x)<br>● Hispanic / Latino / Latina / Latinx<br>● Not Hispanic / Latino / Latina/ Latinx<br>● I don't know<br>● Skip |
| 40a | **FI: Which of the following is your heritage group? Please check all that apply.**<br>**FF: Which of the following is their heritage group? Please check all that apply.**<br><br>● Central American or Central American descent<br>● Cuban or Cuban descent<br>● Dominican or Dominican descent<br>● Mexican or Mexican descent<br>● Puerto Rican or Puerto Rican descent<br>● South American or South American descent<br>● More than one<br>● I don't know<br>● Other (please specify): [text]<br>● Skip |
| 41 | **FI: What is your racial background?** The categories we use may not fully describe you, but they do match those used by the Census Bureau. ***<u>You may choose more than one. (X)</u>***<br>**FF: What is their racial background?** The categories we use may not fully describe you, but they do match those used by the Census Bureau. ***<u>You may choose more than one. (X)</u>***<br>● American Indian or Alaska Native<br>● Asian<br>● Black or African American<br>● Native Hawaiian or Other Pacific Islander<br>● White<br>● I don't know<br>● Skip |
| 42 | **FI: How old are you? (x)**<br>**FF: How old is the person you know? (X)**<br>● Less than 18<br>● 18-30 years old<br>● 30-44 years old<br>● 45-64 years old<br>● Older than 65 years old<br>● I don't know<br>● Skip |
